# Information-Guided Parameter Optimisation for MR Elastography Radiomics

**DOI:** 10.64898/2026.03.17.26348578

**Authors:** Ilies Djebbara, Ziying Yin, Ancuta Ioana Friismose, Frantz Rom Poulsen, Emi Hojo, Jan Saip Aunan-Diop

## Abstract

Mechanical properties of biological tissues vary across spatial scales, yet radiomics typically relies on fixed, heuristic choices for neighbourhood size, kernel geometry, and spectral content - choices that can silently reshape the feature space before any modelling begins. We introduce a label-free, information-theoretic framework for selecting extraction parameters in multi-frequency MRE radiomics. For each configuration θ - neighbourhood radius r, kernel geometry k (sphere or shell), and frequency subset f - we extract a radiomics feature matrix and score it using an objective J(θ) that integrates distributional richness (Shannon entropy), cross-frequency coherence (canonical correlation), inter-feature redundancy (Spearman correlation), and bootstrap stability. We evaluate 121 configurations per tissue in multi-frequency MRE (30-60 Hz) of human brain, liver, and a calibrated phantom, and test robustness using 10,000 Dirichlet-sampled objective weightings. Across tissues, neighbourhood aggregation is consistently preferred over voxel-wise extraction, outperforming the no-neighbourhood baseline in 98.4-100% of weightings. External validation in 100 independent brain scans acquired with a different protocol and wider frequency range (20-90 Hz) confirms a reproducible mesoscopic plateau at r = 3-5 (9-15 mm), with a modal optimum at r = 4; omitting neighbourhood analysis reduces J(θ) by 38% relative to each subject’s optimum. Frequency-subset preferences replicate across datasets, with lower frequencies most frequently selected for brain. By turning ad hoc extraction choices into an outcome-free optimisation step, this framework improves reproducibility, reduces sensitivity to heuristic parameter choices, and generalises across acquisition protocols and imaging sites.

## 1. Background

Mechanical structure in biological tissue is inherently scale dependent. In brain, liver, and engineered gels, relevant microstructural organisation spans from cellular and laminar architecture through mesoscopic boundaries and macroscopic compartments(Sack, 2022). MR elastography (MRE) provides non-invasive access to viscoelastic properties by imaging shear-wave motion and reconstructing the complex shear modulus *G*\* = *G*′ + *iG*″, enabling quantitative maps of stiffness-related and dissipative behaviour(Manduca et al., 2021). Multifrequency MRE further probes dispersion by sampling multiple excitation frequencies, offering complementary sensitivity to wavelength-dependent penetration depth, boundary sharpness, and inversion stability(Klatt et al., 2007; Sack, 2022). These attributes make MRE a promising substrate for radiomics-style feature extraction aimed at characterising tissue state beyond region-of-interest averages(Gillies et al., 2016; Manduca et al., 2021; Sack, 2022).

However, radiomics features are not invariant to upstream extraction choices. First, the neighbourhood used to aggregate local statistics sets the effective analysis scale: small kernels preserve heterogeneity but are noise-sensitive, whereas large kernels stabilise estimates at the cost of blurring anatomical or pathological boundaries(Bernatowicz et al., 2021). This is not only a technical concern: ex vivo rheometry in meningiomas demonstrates substantial intra-tumour viscoelastic heterogeneity, with distinct mechanical phenotypes within the same tumour(Aunan-Diop et al., 2026). Similar phenomena can be observed by varying frequency, measurement time scale and measurable volumes in other tissues, and using other characterization techniques(Reiter et al., 2021). Accordingly, the spatial scale at which mechanical heterogeneity occurs can itself carry clinically useful information and should be preserved rather than averaged away(Aunan-Diop et al., 2026). Second, the excitation-frequency subset used for feature construction determines the spectral information available and mediates a trade-off between penetration, boundary sharpness, and inversion stability in multi-frequency MRE.

For interpretability and reproducibility, these choices matter. In heterogeneous tissue, clinically relevant signal may be encoded in spatial variation rather than region-level averages(Reiter et al., 2021; Streitberger et al., 2020). If the analysis scale or frequency content is chosen heuristically, important structure can be blurred, and downstream models become difficult to reproduce across datasets and protocols. In practice, suboptimal extraction choices may obscure mechanically distinct tissue phenotypes, such as tumour infiltration zones, fibrotic boundaries, or oedema, that are directly relevant to diagnosis, treatment planning, and disease monitoring.

In this study, we use the term information-guided to denote a label-free optimisation strategy that seeks feature spaces with high distributional richness, low redundancy, and robust cross-frequency structure before any supervised modelling is performed. Here, information is understood as observable differences between locations that reduce uncertainty about the underlying tissue structure. The framework is information-guided in the strict sense that it incorporates Shannon entropy as an explicit measure of feature richness, while complementing this with statistical measures of cross-frequency coherence, inter-feature redundancy, and bootstrap stability(Shannon, 1948). Accordingly, the objective is a composite criterion designed to quantify the practical information yield of candidate extraction configurations. We aim to quantify and optimise the information yield of multifrequency MRE radiomics across neighbourhood scales and frequency choices using a reproducible, data-driven selection strategy and verify the strategy on an external dataset.

## 2. Materials and Methods

### 2.1. Problem formulation and objective function

We formalised extraction-parameter selection as a label-free optimisation over configurations *θ* =(*r, s, F*), where *r* is neighbourhood radius, *s* ∈ {none,sphere,shell} denotes kernel geometry, and *F* ⊆ {30,40,50,60} Hz is the excitation-frequency subset with |*F*| ≥ 2: 2. For each *θ*, feature extraction yields a feature matrix *X*(*θ*). We evaluate *X*(*θ*) using an objective function *J*(*θ*) designed to favour feature spaces suitable for unsupervised biomarker discovery, combining: (i) richness *H* (*θ*), defined as mean Shannon entropy across features; (ii) cross-frequency coherence *C* (*θ*), quantified via canonical correlation between frequency-specific feature sets; (iii) redundancy *R*(*θ*), quantified as mean absolute Spearman correlation across feature pairs; and (iv) stability *S*(*θ*), quantified via bootstrap resampling consistency(Hanchuan Peng et al., 2005). Component scores are min-max normalised within tissue to ensure comparability across configurations and combined as:

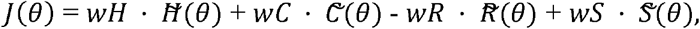

with default equal weights *w*_*H*_=*w*_*C*_=*w*_*R*_=*w*_*s*_=0.25. An explicit baseline configuration *J*(0)=(0, none, *F*) was defined, i.e. no neighbourhood aggregation at the same frequency subset *F*, so that the benefit of neighbourhood context is assessed within the same spectral setting. A complexity penalty P - defined as the equally weighted sum of normalised neighbourhood radius and normalised frequency count, *P* = 0.5 (*r*/*r*_*max*) + 0.5 (|*Ω*|/|*Ω_max*|) - was reported descriptively but excluded from *J*(*θ*) to avoid structural bias toward simpler configurations at the boundary of the parameter space. Robustness to objective weighting was evaluated using a Dirichlet sensitivity analysis (10,000 weight draws over the simplex). The full optimisation was performed over 121 configurations per tissue (11 spatial settings crossed with 11 frequency subsets).

Each component of *J*(*θ*) is designed to capture a distinct and necessary property of a useful radiomics feature space. The four components are defined below alongside the rationale for their sign in the objective.

#### Richness H (positive)

H(θ) measures how much the feature values vary across different parts of the tissue. Higher H means the features capture real spatial differences, which is necessary for them to be informative. H is rewarded in J because useful features should reflect heterogeneity. However, high H alone is not enough, since random noise can also create variation.

#### Cross-frequency coherence C (positive)

C(θ) measures how similar the spatial patterns are across the selected MRE frequencies. If a pattern reflects real tissue structure, it should appear consistently across frequencies, not just in one noisy map. C is therefore rewarded in J. This term is especially sensitive to neighbourhood size, because small amounts of averaging reduce voxel-level noise and make the underlying spatial pattern easier to detect.

#### Redundancy R (negative)

R(θ) measures how much the features overlap with each other. High redundancy means the features are telling us the same thing, so the feature set becomes less useful. R is penalised in J to avoid this. It also prevents the method from favouring overly large neighbourhoods, where too much averaging blurs the tissue and makes different features look increasingly similar.

#### Stability S (positive)

S(θ) measures how reproducible the feature values are when the spatial blocks are resampled. High S means the results are consistent and not overly dependent on exactly which blocks were included. S is rewarded in J because stable features are more likely to reflect real tissue properties.

### 2.2. Data

Two publicly available multi-frequency MRE datasets were used: (i) a development dataset spanning brain, liver, and a calibrated phantom, and (ii) an independent brain dataset acquired with a different protocol and frequency range, used to test reproducibility of the optimisation conclusions.

The development cohort(Feng et al., 2025) consisted of single-subject brain, liver, and phantom MRE were acquired at 3T with four excitation frequencies (30, 40, 50, 60 Hz). Motion was encoded with a spin-echo-based sequence (three motion-encoding directions, four phase offsets). The phantom comprised homogeneous gel compartments (hard gel, soft gel, and background gel), enabling ground-truth separation analyses. The release provides TWENN-reconstructed *G*^*^ maps; to enable frequency-subset evaluation, we computed per-frequency viscoelastic maps from the displacement field using algebraic Helmholtz inversion(Meyer et al., 2022; Oliphant et al., 2001). Dataset characteristics are summarised in Table 1.

**Table 1.**
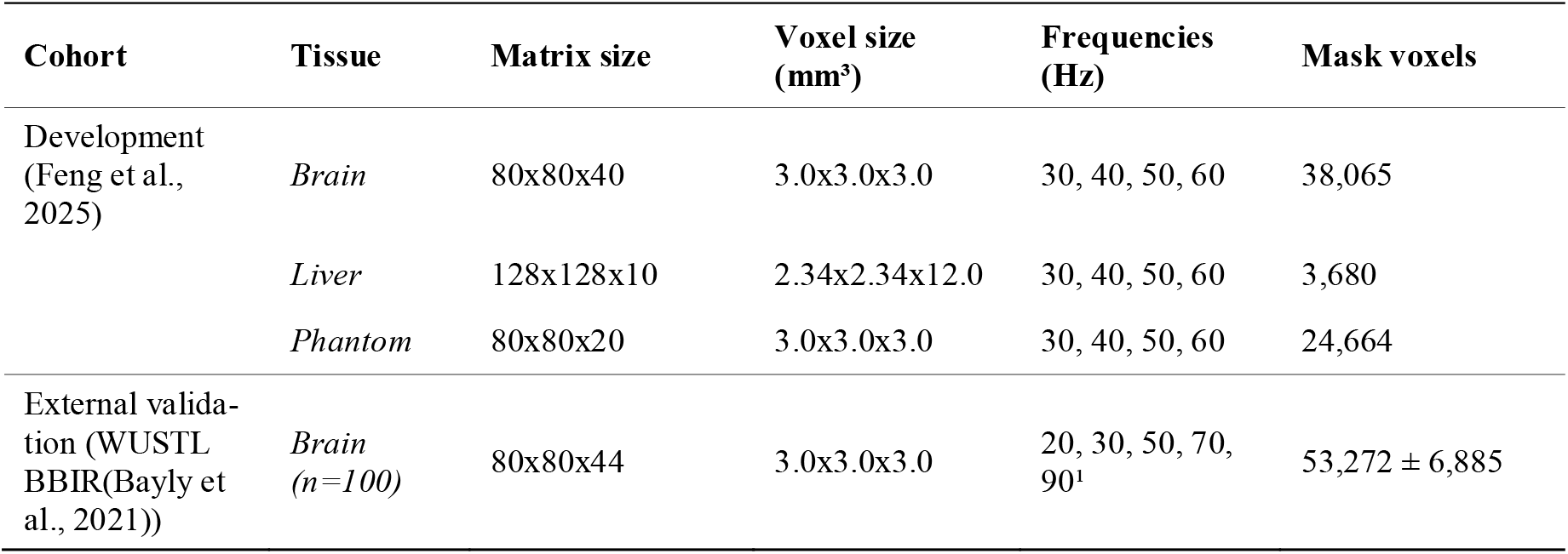
Dataset characteristics. Dataset characteristics for the development cohort (Feng et al., 2025) and the external validation cohort (WUSTL BBIR; Bayly et al., 2021). Per-frequency G′ maps were reconstructed using algebraic Helmholtz inversion (AHI). Mask voxels denote the number of voxels included after tissue segmentation. Liver voxel size reflects 10 mm nominal slices with a 2 mm gap (effective slice thickness 12 mm). The phantom mask includes hard inclusion, soft inclusion, and background gel regions. In external validation, admissible frequencies were restricted by a prespecified AHI quality filter; 90 Hz was excluded in 64% of subjects and 70 Hz in 1%, yielding 3-4 retained frequencies per subject.

| Cohort | Tissue | Matrix size | Voxel size (mm<sup>3</sup>) | Frequencies (Hz) | Mask voxels |
| --- | --- | --- | --- | --- | --- |
| Development (Feng et al., 2025) | *Brain* | 80x80x40 | 3.0x3.0x3.0 | 30, 40, 50, 60 | 38,065 |
|  | *Liver* | 128x128x10 | 2.34x2.34x12.0 | 30, 40, 50, 60 | 3,680 |
|  | *Phantom* | 80x80x20 | 3.0x3.0x3.0 | 30, 40, 50, 60 | 24,664 |
| External validation (WUSTL BBIR (Bayly et al., 2021)) | *Brain (n=100)* | 80x80x44 | 3.0x3.0x3.0 | 20, 30, 50, 70, 90<sup>†</sup> | 53,272 ± 6,885 |

The external validation cohort(Bayly et al., 2021) consisted of 100 independent brain MRE scans acquired at 3T with excitation frequencies 20, 30, 50, 70, and 90 Hz (Table 1). External validation was restricted to brain, as no equivalent multi-subject liver or phantom datasets with compatible acquisition parameters were publicly available. To ensure stable per-frequency AHI reconstructions, admissible frequency subsets were restricted per subject using a prespecified quality filter: frequencies were retained only if the median *G*′ at that frequency was ≤8 ×the subject’s 30 Hz median. This resulted in 3-4 retained frequencies per subject; 90 Hz was excluded in 64% of subjects and 70 Hz in 1% (Table 1). The WUSTL BBIR release provides displacement fields only; no pre-computed stiffness maps are included in the dataset, so per-frequency G′ maps were obtained by applying AHI to the provided displacement fields.

### 2.3 Per-frequency stiffness maps

We computed per-frequency stiffness maps using algebraic Helmholtz inversion (AHI) from the raw displacement field(Meyer et al., 2022; Oliphant et al., 2001). For each frequency *f*, complex displacement was converted to physical displacement using the published motion-encoding efficiencies. The complex shear modulus was estimated as:

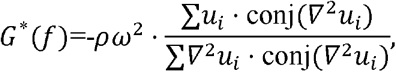

where *ρ* =1000kg/m^3^, *ω=*2*πf*, and *∇*^2^denotes the Laplacian computed with anisotropic voxel spacing. A fixed 3 *x* 3 *x* 3 uniform smoothing filter was applied prior to Laplacian computation and held constant across all configurations(Manduca et al., 2003). The storage modulus *G*′(*f*)=Re[*G*^*^ (*f*)] served as the primary per-frequency feature source. The same fixed 3×3×3 voxel smoothing kernel was applied across all tissues including liver, which has anisotropic voxel spacing (2.34×2.34×12 mm); in physical space this corresponds to approximately 7×7×12 mm, such that the z-dimension is smoothed over a substantially larger extent than the in-plane dimensions. No curl pre-processing was applied to remove longitudinal wave components; AHI was applied directly to the raw displacement field under the standard assumption that shear waves dominate in soft tissue at the employed excitation frequencies. Both the kernel anisotropy for liver and residual longitudinal wave contamination are acknowledged as limitations of the AHI reconstruction. Per-frequency AHI-derived *G*′maps showed the expected increase with excitation frequency and were visually consistent with the TWENN combined reconstruction (Supplementary Fig. S1).

### 2.4 Segmentation and block decomposition

Brain parenchyma was segmented using Otsu thresholding with morphological cleanup (filling, erosion, largest-component selection)(Gonzalez and Woods, 2018; Otsu, 1979). Liver and phantom regions were segmented manually in 3D Slicer(Fedorov A et al., 2012); phantom compartments were labelled as hard gel, soft gel, and background. Each masked volume was decomposed into non-overlapping three-dimensional blocks to provide local spatial replicates for entropy estimation and bootstrap resampling (residual spatial autocorrelation is considered in the limitations). The development dataset was used for method construction; generalisable conclusions are supported by the independent external validation cohort. Blocks were included if ≥ 70% of voxels fell within the mask to ensure block statistics are tissue-dominant (Table 2).

**Table 2.** Block decomposition parameters. Block decomposition parameters for each tissue type. Non-overlapping three-dimensional blocks were used as spatial replicates for entropy estimation and bootstrap resampling. Blocks with <70% mask overlap were excluded. Liver used anisotropic blocks (4 × 4 × 2 voxels) due to the limited number of slices (n = 10). Residual spatial autocorrelation between adjacent blocks is considered in the Limitations.

| Dataset | Block size | Blocks (n) |
| --- | --- | --- |
| *Brain* | 6 x 6 x 6 | 145 |
| *Liver* | 4 x 4 x 2 | 98 |
| *Phantom* | 4 x 4 x 4 | 309 (hard: 37, soft: 29, background: 243) |

### 2.5 Parameter space and feature extraction

The parameter space comprised 121 configurations: 11 spatial settings (baseline *r*=0, and *r* ∈ {1, …,5} *X* {sphere,shell}) crossed with 11 frequency subsets (all subsets of {30, 40, 50,60} Hz with |*F*| ≥ 2: six pairs, four triplets, and one quadruplet). A sphere includes all voxels within Euclidean distance *r*, whereas a shell includes only voxels at distance *r*(±0.5). At *r*=1, both shapes reduce to the six face-adjacent neighbours; a built-in sanity check confirmed identical scores for these configurations. For each configuration, we extracted three feature groups: (i) per-frequency block statistics (8 *x*|*F*|: mean, SD, 10th/50th/90th percentiles, IQR of *G*′, mean *G*′′, and loss ratio *η* = *G*′′/*G*′, the ratio of loss to storage modulus, equivalent to the loss tangent t*an*); (ii) per-frequency neighbourhood statistics (6*x*|*F*|, *r*>0 only: median and IQR of voxel-level neighbourhood mean, SD, and CV); and (iii) cross-frequency features (linear slope and CV across *F*). Total feature dimensionality varied by *θ* and is reported in Table 3.

**Table 3.** Top-ranked configurations by J(θ) Top three configurations ranked by *J*(θ)for each tissue type in the development dataset. 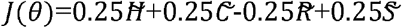, where 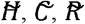, and 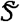 denote min-max normalised richness, coherence, redundancy, and stability, respectively. Configuration notation is *r* [radius] [shape], followed by the frequency subset. A complexity penalty *P*(*θ*) was reported descriptively but excluded from *J*(*θ*)to avoid structural bias toward parameter-space boundaries.

| Rank | Brain | Liver | Phantom |
| --- | --- | --- | --- |
| 1 | r3_shell, 30,40 Hz ($J = 0.517$) | r3_sphere, 30,40 Hz ($J = 0.505$) | r3_shell, 30,40,50,60 Hz ($J = 0.418$) |
| 2 | r3_sphere, 40,50,60 Hz ($J = 0.511$) | r3_shell, 30,40 Hz ($J = 0.503$) | r5_shell, 50,60 Hz ($J = 0.418$) |
| 3 | r3_shell, 40,50,60 Hz ($J = 0.510$) | r2_shell, 30,40 Hz ($J = 0.499$) | r4_sphere, 30,40,50,60 Hz ($J = 0.410$) |

### 2.6 Sensitivity, validation, and feature importance

Robustness to objective weighting was assessed by drawing 10,000 weight vectors from a symmetric Dirichlet distribution (*α*=1) over the 3-simplex (Frigyik BA et al., 2010). Phantom ground-truth validation used leave-one-out LDA classification (accuracy, AUC) and Cohen’s *d* for hard-vs-soft block separation (37 hard, 29 soft). Feature importance was quantified using a Mann-Whitney U-derived AUC and transformed to an importance score(Hanley and McNeil, 1982):

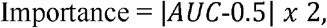

yielding a 0-1 scale where 1 indicates perfect separation and 0 indicates chance-level discrimination.

Additional sensitivity analyses examined the dependence of the spatial optimisation result on the inversion algorithm and on the degree of pre-Laplacian displacement-field smoothing used for AHI; these analyses are reported in Supplementary Section S2.3-S2.4.

### 2.7 External validation

External validation was performed by repeating the full optimisation over *θ*=(*r, s, F*) independently for each subject in the 100-subject brain cohort, using the same feature extraction and objective computation as in the development analyses. Admissible frequency subsets were restricted to the frequencies retained by the prespecified quality filter (2.2). All comparisons were paired within subject across that subject’s evaluated configurations.

To quantify parameter misspecification, the relative loss at configuration *θ*was defined as [*J*(*θ*^∗^)-*J*(*θ*)]/*J*(*θ*^∗^), where *θ*^∗^ denotes the per-subject optimum over the evaluated configuration space. Summary values were computed per subject and averaged across the cohort (Table 5). An overview of all analytical steps is provided in Fig. 1.

**Figure 1.**
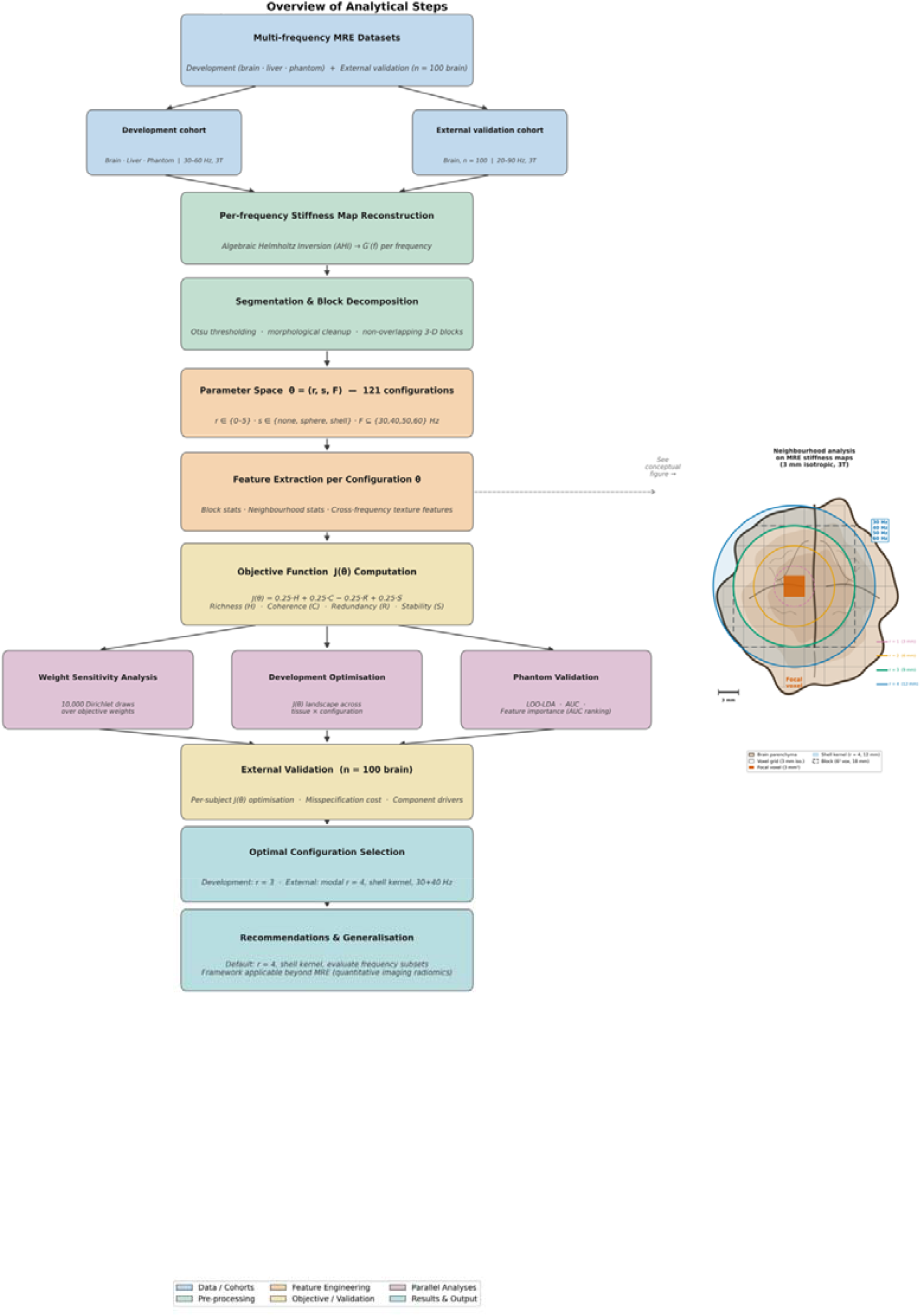
Overview of analytical steps. Multi-frequency MRE data from a development cohort (brain, liver, and phantom; 30-60 Hz, 3T) and an independent external validation cohort (brain, n = 100; 20-90 Hz, 3T) were processed in parallel. Per-frequency stiffness maps (*G*′) were reconstructed using algebraic Helmholtz inversion (AHI), followed by tissue segmentation and three-dimensional block decomposition. Features were extracted across 121 configurations *θ*=(*r, s, F*), defined by neighbourhood radius (*r* ∈ {0, …,5}), kernel geometry (*s* ∈ {none,sphere,shell}), and a frequency subset *F* ⊆ (development: *F* ⊆ {30,40,50,60}Hz; external validation: *F* ⊆ {20,30,50,70,90}Hz after the prespecified quality filter). The label-free objective 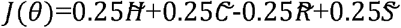 was computed per configuration, combining normalised richness 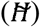, coherence 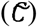, redundancy 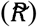, and stability 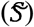. Development analyses comprised landscape optimisation, Dirichlet weight sensitivity (10,000 draws), and phantom ground-truth validation. Optimal configurations were then confirmed in the external cohort by per-subject optimisation and misspecification-cost quantification. Inset: schematic of neighbourhood feature extraction on a representative brain slice; concentric rings indicate radii *r*=1-4, with the shell kernel at *r*=4 (12 mm) highlighted and the focal voxel marked (red square).

## 3. Results

### 3.1 Development dataset: baseline and frequency subset selection

We first quantified the no-neighbourhood baseline *J*(0) and the independent contribution of frequency-subset selection. The baseline achieved high normalised richness (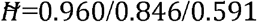 for brain/liver/phantom) but substantially lower cross-frequency coherence 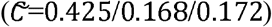 than neighbourhood configurations 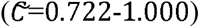. Across 10,000 Dirichlet weight draws, *J*(0) was outperformed in 98.4% (brain), 100% (liver), and 100% (phantom) of trials (Fig. 3C). Frequency-subset selection contributed independently: a subset outperformed all four-frequency configurations in 89.5% (brain), 90.1% (liver), and 74.3% (phantom) of trials, reflecting that the phantom optimum used all four frequencies. The results indicate that both neighbourhood context and spectral choice materially affect information yield, motivating joint optimisation over *θ* =(*r, s, F*).

### 3.2 Development dataset: J(θ) landscape and optimal configurations

In an exploratory analysis on single-subject development data (n=1 per tissue), optimisation landscape was mapped across tissues to identify preferred spatial and spectral settings. Across brain, liver, and phantom, the development data identified *r*=3 as the optimal neighbourhood radius (Table 3; Fig. 2), after excluding the complexity penalty; with the penalty included, optima shifted to *r*=0 (brain) and *r*=4 (phantom). At 3 mm isotropic resolution, *r*=3 corresponds to a 9 mm radius (diameter 7 voxels; 21 mm across); for liver (anisotropic voxels), the corresponding in-plane radius is ∼8 mm. At matched radius, shell kernels outperformed spheres in brain and phantom, whereas liver showed a small sphere advantage (*J* =0.505 vs 0.503). Brain and liver favoured 30+40 Hz, while the phantom favoured all four frequencies; among four-frequency configurations, the optimal configuration placed 6th in brain (*J* =0.506) and 6th in liver (*J* =0.465). Overall, these exploratory findings suggest that the main gains arise from introducing neighbourhood aggregation and selecting informative frequency subsets, with kernel geometry providing secondary but consistent improvements in brain and phantom.

**Figure 2.**
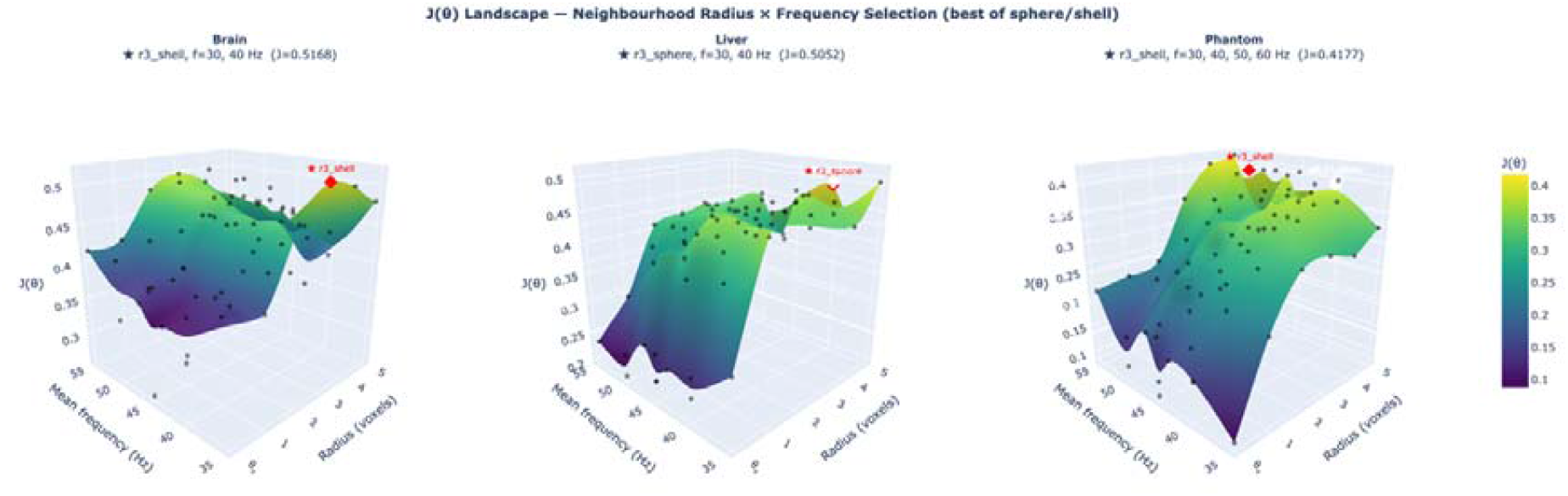
Development landscape. *J*(*θ*) landscape in the development dataset shown as 3D surface plots over neighbourhood radius and the mean frequency of the selected subset, taking the best-performing kernel geometry (sphere or shell) at each point. The colour scale encodes *J*(*θ*). In the single-scan development data, brain and liver exhibit a peak at *r*=3followed by a decline at larger radii, whereas the phantom landscape remains comparatively flat across *r*=3-5. The population-level pattern is evaluated in external validation (Figure 2B).

**Figure 2B.**
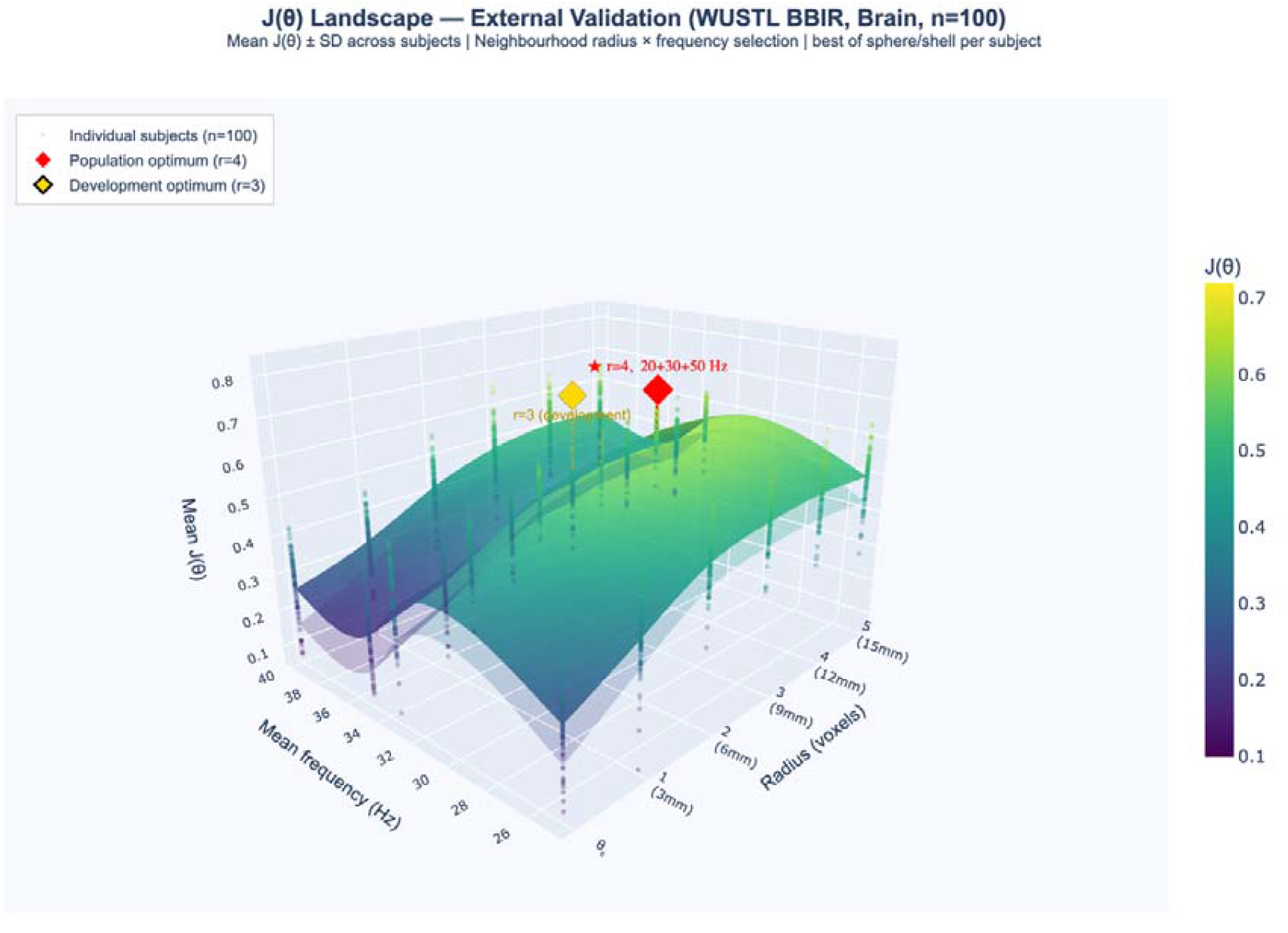
External-validation landscape. J(θ) landscape across the external validation cohort (WUSTL BBIR, brain, n = 100). The upper surface shows the mean *J*(*θ*) across subjects at each (*r, F*) configuration; the translucent lower surface shows mean − 1 SD, with the separation between surfaces reflecting cross-subject variability. Individual subject observations are plotted as points. The red diamond marks the population optimum (*r*=4, 20,30,50 Hz); the gold diamond marks the development optimum (*r*=3) for comparison. The ridge spanning *r*=3-5 indicates a reproducible mesoscopic plateau, whereas the steep descent toward *r*=0 highlights the cost of omitting neighbourhood analysis. Frequency subsets shown are those with universal coverage across all subjects after filtering.

### 3.3 Development dataset: component behaviour and robustness checks

We examined how objective components changed from baseline to optimum and whether conclusions were robust to reasonable implementation choices. Coherence provided the largest baseline-to-optimum gains (Table 4; Fig. 3A): +0.55 (brain), +0.78 (liver), and +0.43 (phantom) in normalised units, while richness peaked at low radii and redundancy decreased with increasing radius (e.g., at *r*=3, 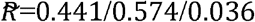 vs 0.767/0.706/0.321 at baseline for brain/liver/phantom). Stability was high overall (brain 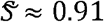; liver 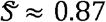; *n*=98 liver blocks). Robustness checks supported the same qualitative conclusions: recomputing coherence with ridge-regularised CCA (*α*=0.1) reduced absolute coherence values uniformly by ∼0.05; redefining redundancy using mean squared (instead of mean absolute) Spearman correlation yielded near-identical configuration rankings (*ρ* =0.999 across 100 external subjects), with the optimal radius identical in 94/100 subjects and the modal optimum unchanged at *r*=4 (Supplementary Analyses S2.1, S2.2). In contrast, removing the redundancy term shifted the optimal radius in 52% of subjects, indicating that redundancy control is a key driver of the selected spatial scale. Further supplementary analyses supported preservation of the r = 3-5 mesoscopic plateau under variation in the pre-Laplacian smoothing kernel and when using k-MDEV inversion instead of AHI (Supplementary Section S2.3-S2.4).

**Figure 3.**
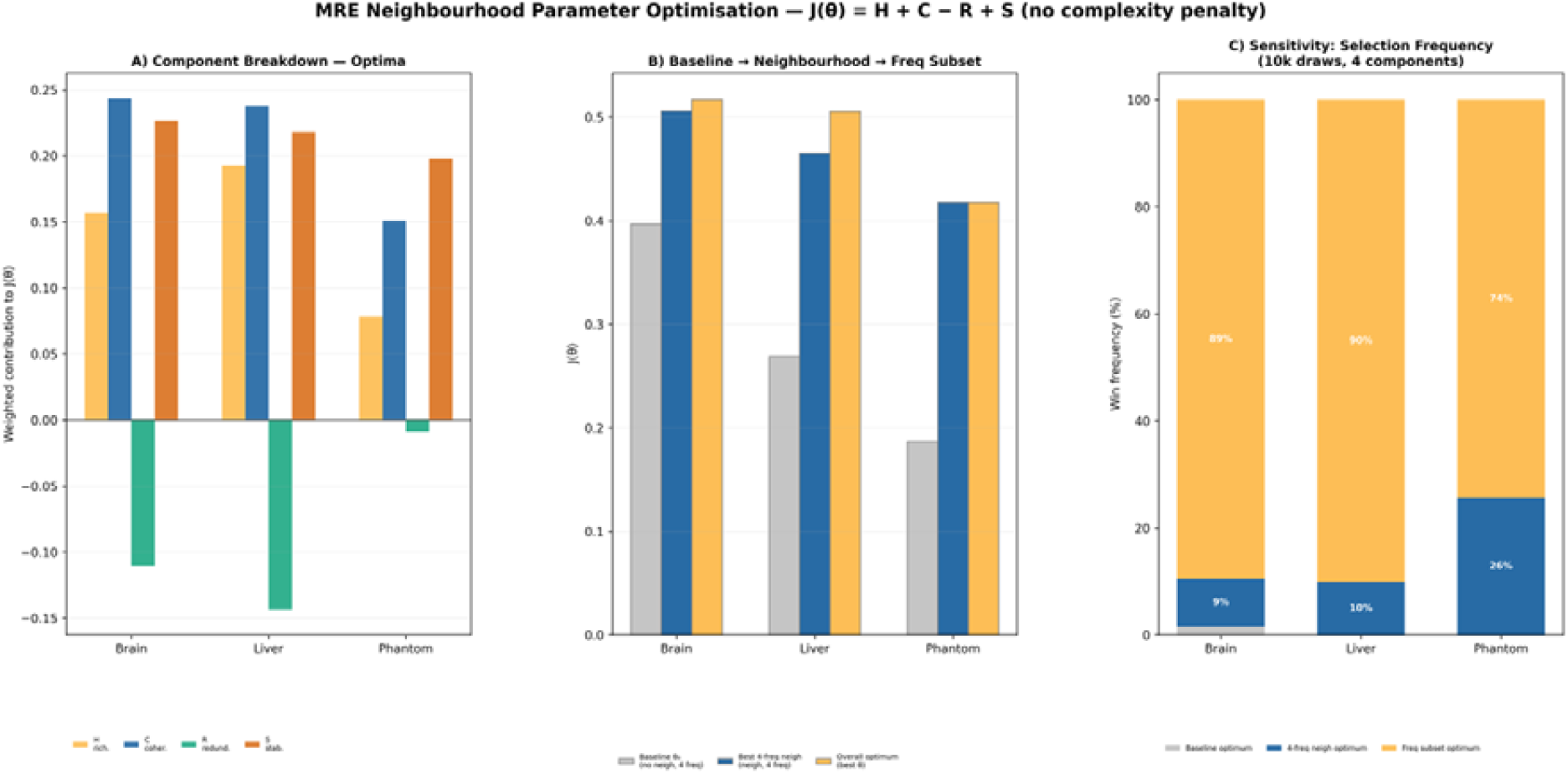
Summary of the optimisation. Summary of the information-theoretic optimisation. (A) Component breakdown at each tissue’s optimal configuration, showing the weighted contributions of richness 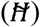, coherence 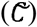, redundancy 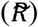, and stability 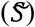 to (*θ*). (B) Progressive improvement from the no-neighbourhood baseline to the best four-frequency neighbourhood configuration, and then to the overall optimum with frequency-subset selection. (C) Selection frequency across 10,000 Dirichlet-sampled objective weightings, partitioned into baseline selections, four-frequency neighbourhood selections, and frequency-subset optima.

**Table 4.** Min-max normalised component values at the optimal configuration. Min-max normalised objective components for the no-neighbourhood baseline and the optimal configuration in the development dataset, reported separately for brain, liver, and phantom. 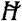 denotes distributional richness (Shannon entropy), 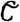 cross-frequency coherence (canonical correlation), 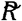 inter-feature redundancy (mean absolute Spearman correlation), and 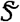 bootstrap stability. The baseline configuration uses no neighbourhood aggregation (*r*=0) with the same frequency subset as the corresponding optimal configuration, so that baseline-to-optimum differences isolate the contribution of neighbourhood aggregation and geometry rather than spectral choice. Higher values of 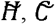, and 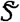 and lower values of 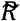 contribute positively to *J*(θ).

| Component | Baseline $\theta \square^1$ | Brain optimum | Liver optimum | Phantom optimum |
| --- | --- | --- | --- | --- |
|  | *(Brain / Liver / Phantom)* |  |  |  |
| $\tilde{H}$ (richness) | 0.960 / 0.846 / 0.591 | 0.628 | 0.771 | 0.312 |
| $\tilde{C}$ (coherence) | 0.425 / 0.168 / 0.172 | 0.974 | 0.951 | 0.604 |
| $\tilde{R}$ (redundancy) | 0.767 / 0.706 / 0.321 | 0.441 | 0.574 | 0.036 |
| $\tilde{S}$ (stability) | 0.968 / 0.766 / 0.306 | 0.907 | 0.872 | 0.791 |

**Table 5.** Cost of parameter misspecification across 100 external validation subjects (WUSTL BBIR) Mean *J*(*θ*) ±SD and misspecification costs for heuristic radius choices computed across all 100 external validation subjects (WUSTL BBIR). Cost vs optimum is reported as absolute *J*(*θ*) units and as percentage of the per-subject optimum. “n with >0.10 *J* lost” counts subjects whose selected radius yields *J*(*θ*) more than 0.10 below their per-subject optimum. Cohen’s *d* is reported relative to the external modal optimum (*r*=4); “-” indicates comparisons not evaluated as primary. “Gain captured” is the fraction of the achievable improvement from *r*=0 to the per-subject optimum recovered at each radius. All comparisons are paired within subject. The misspecification cost is strongly non-linear: the largest gain occurs at the transition from *r*=0to any neighbourhood configuration. Within the mesoscopic plateau (*r* = 3-5), the mean difference between *r*=3 and *r*=4 is +0.019 in *J*(*θ*), driven by reduced redundancy (*ΔR* = -0.009, *p*=0.0002), with no significant differences in coherence (*p*=0.62), richness (*p* =0.91), or stability (*p* = 0.46). The neighbourhood-vs-baseline gap is 11.4x larger than within-plateau variation.

| Configuration | Mean $J(\theta) \pm \text{SD}$ | Cost vs optimum | n with >0.10 $J$ lost | Cohen's $d$ | Gain captured |
| --- | --- | --- | --- | --- | --- |
| $r = 0$ (no neighbourhood) | $0.355 \pm 0.114$ | -0.217 (-38%) | 100/100 | 1.83 | 0% |
| $r = 1$ (minimal heuristic) | $0.403 \pm 0.067$ | -0.169 (-30%) | 87/100 | - | 22% |
| $r = 2$ (common heuristic) | $0.502 \pm 0.048$ | -0.070 (-12%) | 14/100 | - | 68% |
| $r = 3$ (development optimum) | $0.548 \pm 0.041$ | -0.024 (-4%) | 0/100 | - | 89% |
| **$r = 4$ (external modal optimum)** | $0.566 \pm 0.049$ | -0.005 (-0.9%) | 0/100 | Ref. | 97% |

### 3.4 Weight sensitivity

We assessed whether the preferred configurations depended on the equal-weight setting by sampling 10,000 objective weight vectors from a symmetric Dirichlet distribution (Fig. 3A-C). Across tissues, the no-neighbourhood baseline was rarely selected (<1.6% of trials), indicating that neighbourhood aggregation is preferred under a wide range of objective trade-offs. Selection mass concentrated within a narrow spatial regime: the most frequently selected configurations were centred on mesoscopic radii, with tissue-specific differences primarily expressed through frequency subset and kernel geometry rather than large shifts in radius. In brain, *r*3_shell at 30,40 Hz was selected most often (18.0%). In liver, selections favoured sphere kernels at small radii (*r*2_sphere at 30,40 Hz: 20.3%; *r*3_sphere: 7.2%). In the phantom, selections favoured larger radii and higher-frequency subsets (*r*5_shell at 50,60 Hz: 31.7%), with *r*3_shell using all four frequencies second (14.9%). Overall, these results show that the key conclusions are robust to objective weighting: neighbourhood context is consistently preferred over voxel-wise extraction, and the optimisation concentrates on a small radius range while allowing tissue-specific spectral and geometric preferences within that regime.

### 3.5 Phantom validation and feature importance

To link *J*(*θ*) to a ground-truth separation task, we evaluated hard-vs-soft phantom block classification across configurations (Fig. 4). Separation was strong across the configuration space (Cohen’s *d*>1.24, *p* <10^−12^ ), while the baseline achieved the highest leave-one-out AUC (0.895) despite scoring lowest on *J* (θ), indicating that *J*(*θ*)captures information yield rather than task-specific accuracy. In the feature-importance analysis (Fig. 5), neighbourhood-derived variability features ranked highest; for example, the median neighbourhood SD at 60 Hz (f60_neigh_std_med) achieved AUC = 1.000. Stratifying blocks by distance to the hard-soft interface showed that near-boundary blocks (≤3 mm; *n*=34) benefited most from neighbourhood configurations (LOO-LDA AUC = 1.000 vs 0.974 for baseline), whereas far-from-boundary blocks (>3 mm; *n*=32) showed similar performance (0.959 vs 0.951). Together, these results suggest that neighbourhood features preferentially capture boundary-adjacent structure in otherwise homogeneous material.

**Figure 4.**
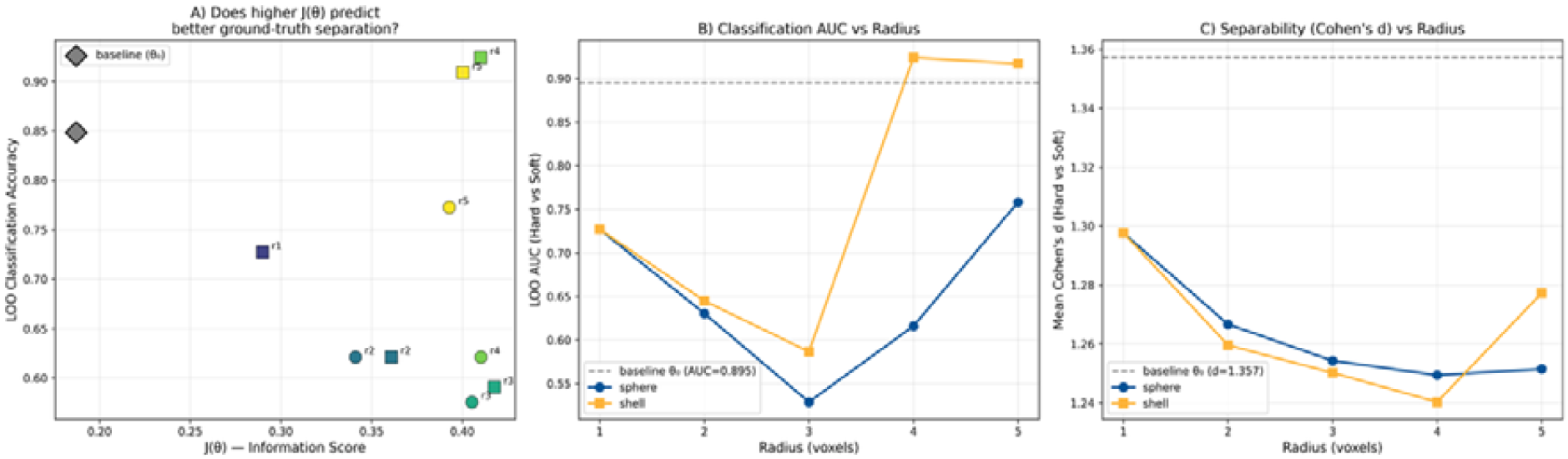
Phantom ground-truth validation. Phantom ground-truth validation. (A) *J*(*θ*)versus leave-one-out (LOO) classification accuracy for hard-versus-soft block discrimination, highlighting that the voxel-wise baseline (grey diamond) can achieve high task accuracy despite low *J*(*θ*). (B) LOO AUC versus neighbourhood radius, with baseline AUC shown as a dashed line. (C) Cohen’s *d* versus radius for hard-versus-soft separation.

**Figure 5.**
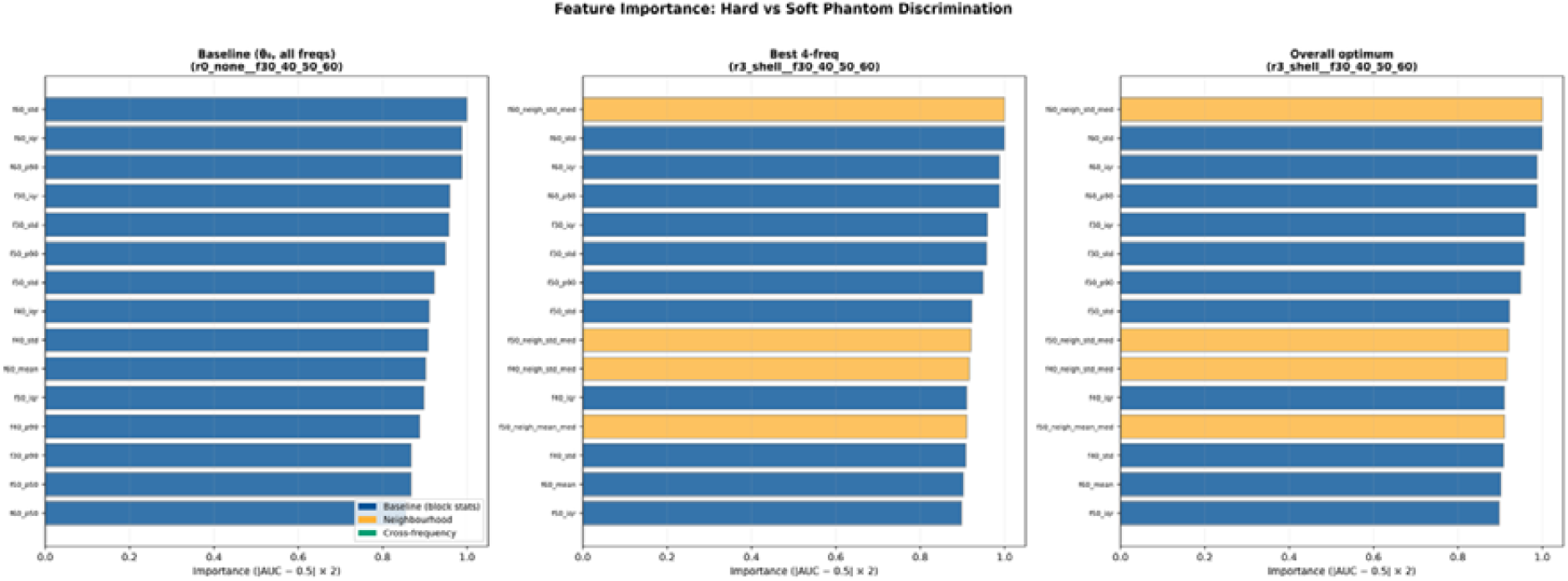
Feature importance in the phantom. Feature importance for phantom hard-versus-soft discrimination. Horizontal bar charts show the top 15 features ranked by univariate AUC for two configurations: the voxel-wise baseline ***θ*_*0*_** (left) and the best neighbourhood configuration (right). Bars are grouped by feature class (block-level statistics, neighbourhood features, and cross-frequency features). The neighbourhood feature f60_neigh_std_med (median neighbourhood SD at 60 Hz) ranks highest when spatial context is included..

### 3.6 External validation: cost of parameter misspecification

We next quantified the practical cost of common heuristic radius choices in the independent 100-subject cohort (Table 5), relative to each subject’s optimum. Omitting neighbourhood analysis (*r*=0) reduced *J* (*θ*) by a mean of 0.217 units (38% of the per-subject optimum), and all subjects lost more than 0.05 *J* relative to their optimum (100/100; paired *t*-test: *d*=1.83, *p*<10^−33^ ). The minimal heuristic (*r*=1) captured 22% of the achievable gain over the *r*=0 baseline, whereas the common heuristic (*r*=2) captured 68% and left 12% of the optimal yield unrealised in 71% of subjects. Misspecification costs were strongly non-linear: most improvement occurred at the transition from *r*=0 to any neighbourhood configuration, with diminishing returns beyond *r*=2. Within the *r*=3-5 plateau, the mean difference between *r*=3 and *r*=4was +0.019 }, driven by lower redundancy (*ΔR*=-0.009, *p*=0.0002), with no significant differences in coherence, richness, or stability; the neighbourhood-vs-baseline gap was 11.4x larger than within-plateau variation. Overall, these results show that the main risk is omitting neighbourhood context, whereas fine-grained tuning within the plateau yields smaller but measurable gains.

### 3.7 External validation: optimal configurations and component drivers

Finally, we characterised the preferred configurations and their drivers in external validation. Neighbourhood analysis was preferred in all 100 subjects. The mean gain from *r*=0 to the per-subject optimum was +0.217 *J*(+54% relative), with a paired effect size of *d*=1.83 (*p*<10^-33^ ); gains ranged from +0.07 to +0.68. The modal optimal radius was *r*=4 (63%), followed by *r*=5 (25%) and *r*=3 (12%); radii *r*=0-2 were not selected (Fig. 2B). Pairwise comparisons within the plateau were statistically significant (*r*=4 vs *r*=3: +0.019, *p*<0.0001; *r*=5vs *r*=3: +0.012, *p*<0.0001), but with smaller effects than the *r*=0 versus optimum comparison (*d*=0.73 vs 1.83). The preference for *r*=4 over *r*=3 was attributable to a modest reduction in redundancy (*ΔR*=-0.009, *p=*0.0002), with no detectable differences in coherence, richness, or stability. Frequency-subset preferences mirrored the development results, with 20, 30, and 50 Hz selected most frequently (62% of subjects), and shell geometry selected in 78% of subjects. Together, these findings indicate a reproducible mesoscopic plateau at *r*=3-5 with a modal optimum at *r*=4, and show that within-plateau preferences are driven primarily by redundancy rather than coherence, richness, or stability.

## 4. Discussion

We introduce a label-free approach to optimise neighbourhood radius and frequency selection in multi-frequency MRE radiomics, addressing a key barrier to reproducibility: upstream extraction settings that reshape the feature space before downstream analysis. Across development data and robustness analyses, neighbourhood aggregation was consistently preferred over voxel-wise extraction, and external validation in an independent 100-subject brain cohort revealed a reproducible mesoscopic plateau in the preferred radius (***r*** = **3*-*5**). The dominant gain came from moving from ***r* = 0** to any neighbourhood configuration, whereas differences within the plateau were comparatively small. Practically, *J*(***θ***) provides an outcome-free pre-modelling step that can be applied before any supervised endpoint is defined, reducing the risk that downstream conclusions reflect arbitrary upstream extraction choices.

### 4.1 Spatial optimisation and tissue-dependent convergence

Neighbourhood aggregation seems to impose a controlled trade-off: larger kernels stabilise local statistics and suppress voxel-level noise, but can blur across anatomical boundaries and reduce local heterogeneity(Bernatowicz et al., 2021). In brain external validation, the preferred radii clustered within a mesoscopic range (*r*=3-5), consistent with kernels that are large enough to improve statistical stability while still respecting dominant parenchymal boundaries at the acquisition resolution. The phantom landscape supports this interpretation: in a largely homogeneous material with sparse internal boundaries, larger radii are less penalised, yielding a flatter optimum region(Okamoto et al., 2011). In brain, performance can decline at larger radii when kernels increasingly mix across grey-white transitions and vascular structures, reducing heterogeneity and increasing redundancy(Sack, 2022). Together, these observations suggest that the information-preferred radius is not an arbitrary choice, but a reproducible analysis parameter that should be reported explicitly in MRE radiomics pipelines.

### 4.2 Frequency selection is a first-order design choice

A key result is that frequency selection materially changes the information yield of multifrequency MRE radiomics: in brain and liver, optimised frequency subsets were preferred over using the full acquired set under most objective weightings. This is important because frequency content is often treated as a fixed property of the acquisition rather than an explicit modelling choice, yet it determines the spectral representation from which features are constructed and therefore the effective feature space(Hiscox et al., 2016; Manduca et al., 2021). Practically, two studies can compute identical radiomics features and still obtain meaningfully different feature spaces if they include different frequency subsets, reducing meaningful clinical information and reproducibility across cohorts(Duron et al., 2019; Traverso et al., 2018).

Mechanistically, multi-frequency MRE involves a trade-off across excitation frequencies between wave propagation/penetration, boundary sharpness, and inversion stability; selecting a subset can therefore increase cross-frequency coherence and stability while limiting redundancy in the resulting feature matrix(Sack, 2022). The phantom’s different spectral preference is consistent with a setting dominated by engineered boundaries rather than heterogeneous biological microstructure (Feng et al., 2025). Methodologically, these findings motivate treating frequency selection as a reportable parameter - the *F* in *θ* = (*r, s, F*) - and reevaluating it when acquisition protocols or cohorts change, rather than assuming maximal frequency inclusion is always optimal.

### 4.3 J(θ) optimises information capacity, not task-specific accuracy

A central property of *J*(*θ*) is that it requires no external labels, making it applicable where conventional supervised feature selection cannot operate. The phantom validation illustrates why this distinction matters: the voxel-wise baseline achieved high hard-vs-soft discrimination despite scoring lowest on *J* (*θ*), because a single block-level contrast (mean *G*′) is already sufficient in a two-compartment phantom. Neighbourhood features can partially smooth across the boundary and therefore need not improve a task that is dominated by a single strong contrast. Stratifying blocks by distance to the hard-soft interface supports this interpretation: neighbourhood configurations provided the clearest advantage near boundaries, where spatial contrast carries the discriminating signal. In contrast, stratification provided less advantage in homogeneous interiors. In biological tissue, by contrast, clinically relevant signal is often encoded in distributed heterogeneity rather than a single compartment mean(Reiter et al., 2021; Streitberger et al., 2020); in such settings, *J*(*θ*) provides an outcome-free way to prioritise extraction configurations before downstream modelling. When labels are available, supervised methods should complement *J*(*θ*); when they are not, *J*(*θ*) avoids the circularity of tuning extraction parameters on the same labels later used for model training(Demircioğlu, 2021).

### 4.4 Choice of inversion algorithm

The Feng et al. dataset provides only a combined TWENN reconstruction; we therefore applied algebraic Helmholtz inversion (AHI) independently at each excitation frequency to obtain per-frequency *G*′ maps. AHI was chosen for transparency, reproducibility, and wide availability(Meyer et al., 2022; Oliphant et al., 2001). Its primary limitation is noise sensitivity: estimating *G*^*^ from local Laplacian operators amplifies measurement noise, particularly at higher excitation frequencies where wavelengths are shorter and spatial gradients steeper(Oliphant et al., 2001). Learned inversion approaches can reduce this noise floor by incorporating spatial priors trained across large datasets, potentially increasing absolute baseline coherence(Ma et al., 2023). Whether that would change the neighbourhood-driven pattern observed here is unknown; however, because AHI noise acts consistently across configurations, the relative ordering of configurations, and thus the preference for neighbourhood aggregation, is unlikely to be fundamentally altered. Consistent with this interpretation, a supplementary sensitivity analysis using k-MDEV in the external validation cohort reproduced the same mesoscopic plateau and modal optimum, despite differences in absolute stiffness scaling between methods; as noted in Supplementary Section S2.3, the two pipelines differ in preprocessing (directional filtering, masking), so absolute G′ values are not directly comparable across methods.

### 4.5 Biomechanical and clinical relevance

The parameter vector *θ* = (*r, s, F*)is a biomechanical modelling choice. Neighbourhood radius sets the spatial scale at which viscoelastic heterogeneity is sampled: if too small, extracted features become noise-dominated; if too large, anatomically or pathologically distinct compartments are averaged together, attenuating contrasts that carry diagnostic or biological meaning. For example, rheometry studies of meningiomas report marked intratumour mechanical heterogeneity, underscoring that clinically relevant signal can reside at sub-lesion scales(Aunan-Diop et al., 2026, 2023). Similar observations have been made in malignant brain tumors and liver(Reiter et al., 2021). Frequency subset selection carries analogous weight, because excitation frequency shapes wavelength-dependent penetration, boundary sharpness, and the balance between elastic and viscous sensitivity in MRE dispersion. This is observed, for example, in MRE studies of the ageing brain, where varying the excitation frequency and the scale of analysis (whole-brain versus regional) affect the measurement outcome(Schattenfroh et al., 2026). A distinct, but related phenomenon is observed when varying measurement time scale(Su et al., 2023). From a clinical standpoint, many questions are inherently scale-specific - for example, boundary stiffness gradients relevant to tumour interface characterisation or compartmental shifts accompanying fibrotic remodelling. In such settings, *θ d*irectly determines whether the radiomics feature space preserves the underlying biomechanical structure at the scale of interest or obscures it through over-smoothing or spectrally suboptimal aggregation. Our results therefore motivate treating *θ* as a reportable methodological parameter: explicitly documenting neighbourhood radius, kernel geometry, and the included frequency subset alongside IBSI-compliant feature definitions improves interpretability and reproducibility across cohorts and protocols (Bernatowicz et al., 2021; Traverso et al., 2018; Zwanenburg A et al., 2020).

The framework may also have broader translational applications. For example, in tumour datasets with paired ex vivo mechanical measurements, such as meningioma cohorts with rheometer-derived stiffness, the same optimisation strategy could be extended to favour feature-extraction settings that are not only rich, stable, and non-redundant, but also most informative about the external mechanical reference(Aunan-Diop et al., 2026). If tumour samples have matching rheometer measurements of storage modulus, the framework could be used to identify the neighbourhood scale, frequency subset, or feature set that best captures imaging differences associated with firmer versus softer tissue. In that context, information would be defined by the extent to which imaging-derived differences reduce uncertainty about underlying tissue mechanics in a clinical context, thereby offering a principled bridge between in vivo MRE and ex vivo biomechanical characterisation.

### 4.6 Limitations

The development analyses rely on block-wise spatial replicates within single scans rather than independent subjects, so local spatial autocorrelation can inflate apparent stability and reduce effective sample size; the key conclusions are therefore anchored in the independent 100-subject external validation for brain. Liver and phantom optimisation conclusions would benefit from multi-subject validation. We used AHI to enable transparent per-frequency reconstruction, but inversion noise and the fixed pre-Laplacian smoothing step may influence absolute component values; however, comparisons were paired within-subject/configuration, so relative rankings are expected to be more robust than raw scores. The current framework treats each tissue as mechanically isotropic; extending the optimisation to account for anisotropic tissues or to stratify by sub-compartments may require additional methodological development. Finally, several fixed pipeline choices (e.g., smoothing and block inclusion threshold) were not jointly optimised with ***θ*** and could be incorporated in future extensions of the framework. Although the exact point optimum showed modest sensitivity to implementation choices, supplementary analyses indicated that the broader plateau result was stable across both inversion method and smoothing-kernel settings (Supplementary Section S2.3-S2.4).

## 5. Conclusion

Neighbourhood radius, kernel geometry, and frequency subset directly determine the information yield of multi-frequency MRE radiomics feature spaces, yet are often treated as implementation details. In external validation (n = 100), omitting neighbourhood analysis (*r*=*0*) reduced *J*(*θ*) by 0.217 units on average (38% of each subject’s optimum; Cohen’s *d*=1.83), demonstrating that the dominant gain arises from introducing spatial context. A label-free objective, *J*(*θ*), enabled principled selection of *θ*=(*r, s, F*)and reproduced across datasets and protocols.

Across three tissues in development data and in all 100 external subjects, neighbourhood configurations were preferred over the no-neighbourhood baseline under 10,000 Dirichlet-sampled objective weightings, with gains primarily associated with increased cross-frequency coherence. In brain MRE at 3 mm isotropic resolution, the information-preferred spatial scale formed a mesoscopic plateau at *r*=3-5 (9-15 mm), with *r*=4 as the modal optimum; the neighbourhood-vs-baseline effect was 11.4x larger than variation within the plateau.

Pragmatically, these findings support treating *θ* as a primary methodological parameter that should be reported explicitly. For brain data at comparable resolution, *r*=4 with a shell kernel is a reasonable starting point within the plateau, but *θ* should be re-evaluated when acquisition protocols, voxel geometry, tissues, or cohorts differ. More generally, *J*(θ) provides an outcome-free way to reduce scale-dependent variability before feature computation and can be applied to other quantitative imaging settings where features depend on neighbourhood scale and multiple acquisition parameters.

## Data availability

All MRE data are publicly available from (Feng et al., 2025) and Brain Biomechanics Imaging Repository (http://www.nitrc.org/projects/bbir). Access to BBIR data, and data derived from BBIR data, is governed by the BBIR data use agreement. The analytical pipeline is available from the corresponding author upon reasonable request.

## Supplementary Materials

**Supplementary Figure S1.**
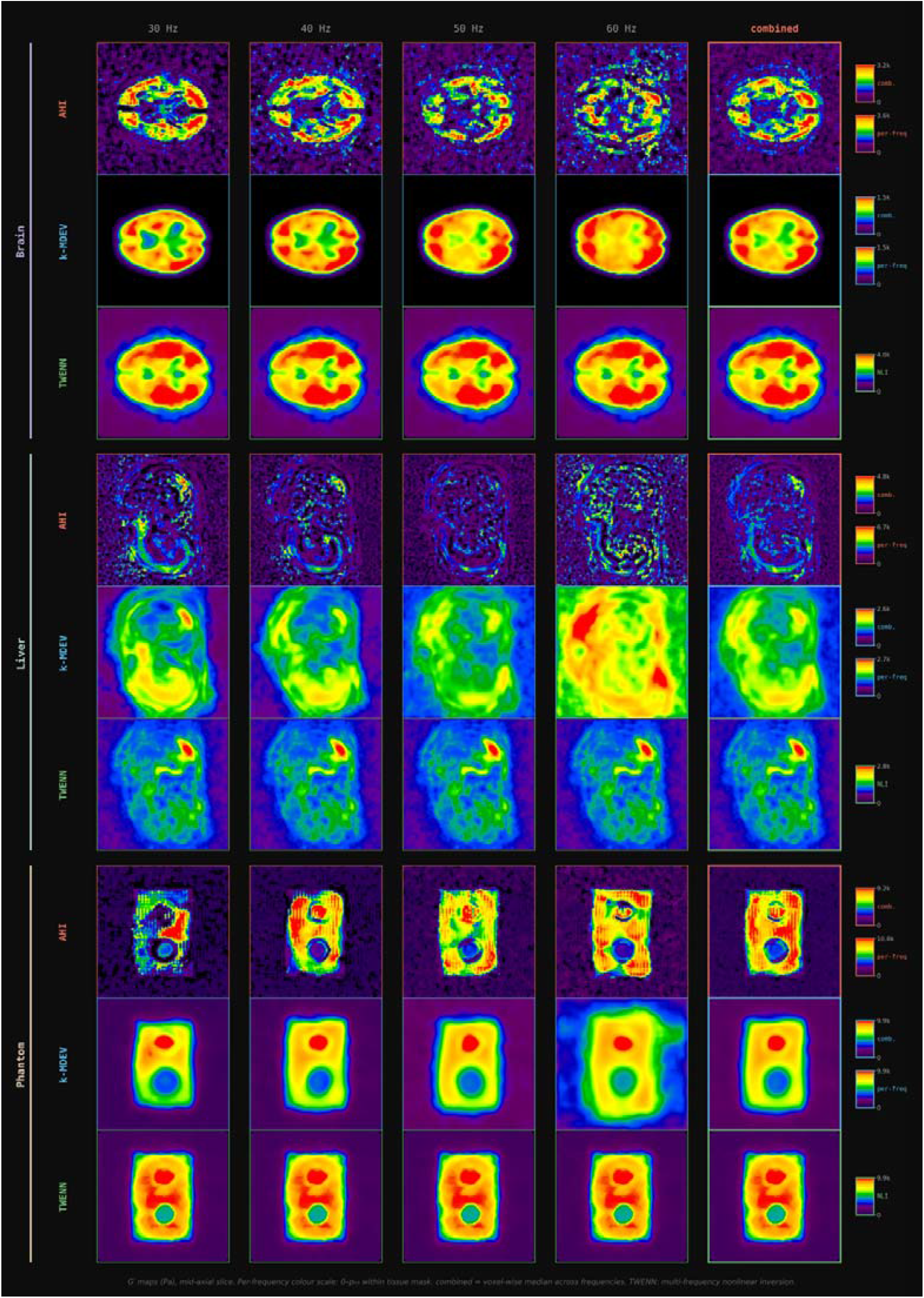
Per-frequency G′ maps for three inversion methods across brain, liver, and phantom. Each panel shows the mid-axial slice at 30, 40, 50, and 60 Hz, alongside a voxel-wise median composite across all four frequencies (combined). Methods shown are AHI (algebraic Helmholtz inversion), k-MDEV (k-space multi-directional elasticity), and TWENN (multi-frequency nonlinear inversion; combined reconstruction only). Colour scales are independent per method and tissue, anchored to the 95th percentile within the tissue mask; brightness therefore reflects relative contrast within each method rather than absolute stiffness comparisons across methods. AHI maps show progressive noise amplification at higher frequencies, particularly in brain (60 Hz) and phantom, consistent with Laplacian instability at shorter wavelengths. k-MDEV preserves anatomical coherence across all frequencies due to directional wavenumber filtering. In the phantom, AHI exhibits banding artefacts from wave interference, whereas k-MDEV clearly resolves the hard and soft inclusions. TWENN and k-MDEV show broadly comparable spatial patterns, particularly in brain and phantom.

## Supplementary analyses

### S2.1 Redundancy operationalisation sensitivity

R(θ) was defined as mean absolute Spearman correlation across all feature pairs. We compared this to a squared alternative (mean ρ^2^), which places greater weight on strongly correlated pairs. Mean Spearman rank correlation between J(θ) scores under the two operationalisation was ρ = 0.999 across n = 100 external validation subjects, with the optimal radius identical in 94/100 subjects and the modal optimum unchanged at r = 4. Removing R from J(θ) entirely - redistributing its weight equally across H, C, and S - produced an identical optimal radius in only 48/100 subjects, with several subjects selecting boundary solutions (r = 0 or r = 2), consistent with coherence dominating an unconstrained objective. The redundancy component is therefore load-bearing: its specific operationalisation does not materially affect results, but its presence is necessary for interior plateau solutions.

### S2.2 Richness component weighting sensitivity

We varied w_H from 0.00 to 0.50, redistributing remaining weight equally across C, R, and S (equal-weight operational setting: w_H = 0.25). Configuration rankings relative to equal weighting, assessed via mean Spearman ρ across n = 100 subjects, were 0.96 (w_H = 0.125) and 0.95 (w_H = 0.375), indicating near-identical rankings under moderate perturbation. The mesoscopic plateau (r = 3-5) contained the optimal radius for 100/100 subjects at w_H = 0.125-0.375. At w_H = 0.000, the modal optimum shifted to r = 5 in 88% of subjects, reflecting the monotonic coherence advantage of large radii without an entropy penalty; at w_H = 0.500, 21/100 subjects selected r = 0, consistent with the entropy advantage of the no-neighbourhood baseline under high richness weighting. Crucially, no subject selected r = 0 for any w_H ≤ 0.375, confirming that the neighbourhood-vs-baseline finding is preserved across all moderate weight settings.

### S2.3 Inversion algorithm sensitivity

This analysis is intended as a sensitivity check for the optimisation result, not a direct comparison between inversion algorithms: AHI and k-MDEV differ substantially in their preprocessing pipelines — k-MDEV applies directional wavenumber filtering across a Fibonacci sphere of orientations before stiffness estimation, whereas AHI operates directly on the raw displacement Laplacian without directional filtering or equivalent masking. Absolute G′ estimates therefore reflect both the inversion mathematics and these pipeline-level differences, and should not be interpreted as a head-to-head performance comparison. To examine the sensitivity of the spatial optimisation result to inversion method, we repeated the full J(θ) optimisation in all 100 external validation subjects using k-space multi-directional elasticity (k-MDEV) inversion instead of AHI. k-MDEV estimates stiffness using directional wave-number filtering across a Fibonacci sphere of orientations and derives stiffness from the ratio of displacement energy to gradient energy in the filtered field, thereby avoiding the pointwise Laplacian ratio used in AHI. Despite these differences in absolute stiffness estimates, the optimisation result was highly similar across methods: under k-MDEV, the optimal radius was r = 3 in 12 subjects, r = 4 in 64, and r = 5 in 24, compared with 12, 63, and 25 subjects, respectively, under AHI. All 100 subjects fell within the same plateau range (r = 3-5) under both inversions, and r = 4 was the modal optimum in both cases. The mean J(θ) profile also peaked at r = 4 for both methods, although k-MDEV yielded uniformly higher absolute J values across r = 2-5. Frequency-subset preferences differed modestly between methods (AHI: 20,30,50 Hz in 62% of subjects; k-MDEV: 50,70 Hz in 40% and 30,50 Hz in 34%), suggesting some method-dependent redistribution of frequency contributions. Overall, these findings support preservation of the main spatial result across inversion methods, namely a mesoscopic plateau spanning r = 3-5 with a modal optimum at r = 4.

### S2.4 Pre-Laplacian smoothing kernel sensitivity

AHI requires pre-smoothing of the displacement field before Laplacian-based stiffness estimation to mitigate voxel-level noise; the default setting used a 3×3×3 uniform filter (ks = 3). As a sensitivity analysis, we repeated the development-dataset J(θ) optimisation using a 5×5×5 kernel (ks = 5). Under ks = 5, the optimal radius shifted from r = 3 to r = 4 in brain and liver, and from r = 3 to r = 5 in phantom. Despite these shifts in the point optimum, all solutions remained within the same plateau range (r = 3-5), and the plateau boundaries were unchanged. The tendency towards larger optimal radii with heavier smoothing is consistent with reduced high-frequency noise increasing the stability of neighbourhood-derived features at somewhat larger spatial scales. In brain and liver, the ks = 5 optimum of r = 4 also aligned with the modal optimum observed in the external validation cohort. Together, these findings suggest that while the exact point optimum may vary modestly with smoothing choice, the principal result - a stable mesoscopic plateau spanning r = 3-5 - is preserved

